# Magnetic Resonance Imaging Radiomics-Based Model to Identify the Pathological Features and Lymph Node Metastasis in Rectal Cancer

**DOI:** 10.1101/2022.05.02.22274247

**Authors:** Peng Chen, Deying Wen, Libin Huang, Jingjing Ding, Wenming Yang, Jiayu Sun, Lie Yang, Zongguang Zhou

**Author notes:** ***Correspondence to:*** Lie Yang. Department of Gastrointestinal Surgery, West China Hospital, Sichuan University, No. 37 Guoxue Lane, Chengdu 610041, China.; <u>Jiayu Sun.</u> Department of Radiology, West China Hospital, Sichuan University, No. 37 Guoxue Lane, Chengdu 610041, China. These authors have contributed equally to this work and share first authorship (Peng Chen, Deying Wen).

## Abstract

**Background:** Pathological features and lymph node staging plays an important role in treatment decision-making. Yet, the preoperative accurate prediction of pathological features and lymph node metastasis (LNM) is challenging.

**Objective:** We aimed to investigate the value of MRI-based radiomics in predicting the pathological features and lymph node metastasis in rectal cancer.

**Methods:** In this prospective study, a total of 37 patients diagnosed with histologically confirmed rectal cancer who underwent pelvic 3.0T magnetic resonance imaging (MRI) were enrolled. MRI images of both the primary tumor alongside the lymph nodes and specimens were performed with a node-to-node match and labeling. The correlation analysis, least absolute shrinkage, and selection operator (LASSO) logistic regression (LR), backward stepwise LR were mainly used for radiomics feature selection and modeling. The univariate and multivariate backward stepwise LR were used for preoperative clinical predictors selection and modeling.

**Results:** A total of 487 lymph nodes including 39 metastatic lymph nodes and 11 tumor deposits were harvested from 37 patients. The texture features of the primary tumors could successfully predict tumor differentiation using a well-established model (area under the curve (AUC) = 0.798). Sixty-nine matched lymph nodes were randomly divided into a training cohort (n = 39) and a validation cohort (n = 30).

Three independent risk factors were obtained from 56 texture parameters closely related to LNM. A prediction model was then successfully developed, which provided AUC values of 0.846 and 0.733 in the training and test cohort, respectively. Further, tumor deposits produced a higher radiomics score (Rad-score) compared with LNM (*P* = 0.042).

**Conclusion:** The study provides two non-invasive and quantitative methods, which respectively predict the tumor differentiation and regional LNM for rectal cancer preoperatively. Ultimately, these are favorable when producing treatment protocols for rectal cancer patients.

## INTRODUCTION

Colorectal cancer is both the 3rd most frequently diagnosed cancer and the 3rd leading cause of cancer death worldwide(1). Moreover, rectal cancers account for approximately 40% and more than 44,850 new cases will be diagnosed in 2022(1, 2). Preoperative staging is routine in the treatment plan decision, while the detection of perirectal LNM is critical for accurate staging. Patients presenting with LNM should receive neoadjuvant therapy followed by radical surgery(3, 4). Preoperative evaluation of LNM is dependent upon imaging methods, predominantly computed tomography (CT), MRI or ultrasound scans. However, these methods remain descriptive approaches that rely on the morphological features of lymph nodes for the diagnosis, which confines the accuracy of preoperative staging and thus may lead to a relatively incorrect treatment plan(5, 6).

Recently, the application of high-resolution MRI has improved the accuracy of local staging. However, the diagnosis of LNM remains unreliable since it depends on the analysis of morphological features, such as bigger node size, irregular borders, and heterogeneous signals(5). Studies have shown that node size was a poor predictor of nodal status because the size of normal or reactive lymph nodes was equal to or greater than that of positive lymph nodes(7, 8). Gröne et al. reported that combining the features of node border and signal did not improve the diagnosis accuracy compared to size criteria, even with high-resolution MRI (76.7% vs 68.3%)(5). Therefore, the descriptive method is fairly unreliable and there is an urgent requirement to establish a quantitative method that improves the accuracy of LNM evaluations.

Radiomics is a recently developed quantitative technique that can potentially transform original monochrome images into quantitative features using high-throughput extraction methods(9, 10). Studies have shown that radiomic features are closely linked to patients’ prognosis and underlying genomic phenotyping across a range of cancers(11-13). Indeed, Huang et al. and Xu et al. have developed predictive models in rectal cancers based on the radiomics nomogram to predict LNM with retrospective data from CT and MRI scans, respectively(14, 15). However, the retrospective characteristics employed greatly limited the reliability of the results. Besides, their studies did not contain any node-to-node comparisons between the radiomics and surgical lymph node histopathology. A prospective study reported that the chemical shift effect is a reliable predictor for differentiating benign from metastatic nodes using high-resolution MRI with node-for-node matched histopathological validation. However, it is difficult to determine the nodal status with only a single predictor in this study. Thus, the MRI radiomics analysis of lymph nodes could provide a more accurate assessment of the nodal status(16).

Currently, there is an absence of prospective study to establish a reliable quantitative model using MRI radiomics for the accurate prediction of pathological features and LNM. In the present study, we prospectively develop and validate MRI radiomic models that predict pathological features and further we adopt the node-to-node method to develop and internally validate an MRI-Based model of LNM prediction in patients with rectal cancer.

## METHODS

### Patient Cohort and Data Collection

All procedures performed in this study involving human participants were in accordance with the Declaration of Helsinki. The involvement of human participants in this study was approved by the Biomedical Ethics Committee of AA Hospital and informed consent was acquired from all patients. From January 2018 to March 2019, the patients diagnosed with rectal cancer and hospitalized for potentially curative surgery were prospectively collected in the gastrointestinal surgery department of the AA Hospital of AA University. The inclusion criteria included patients: (a) with primary rectal adenocarcinoma (including mucinous adenocarcinoma) confirmed by colonoscopy biopsy; (b) requiring radical surgery; (c) undergoing a high-spatial-resolution MRI performed at our institution to a high-quality; (d) MRI examination had been performed fewer than 2 weeks before surgical resection. The exclusion criteria included patients: (a) who had undertaken any preoperative therapies (radiotherapy, chemotherapy, or chemoradiotherapy); (b) possessing a secondary or relapsed rectal cancer. Following the application of the exclusion criterion, 37 patients were selected for inclusion in this study. The flowchart of this study is shown in Figure 1.

**FIGURE 1.**
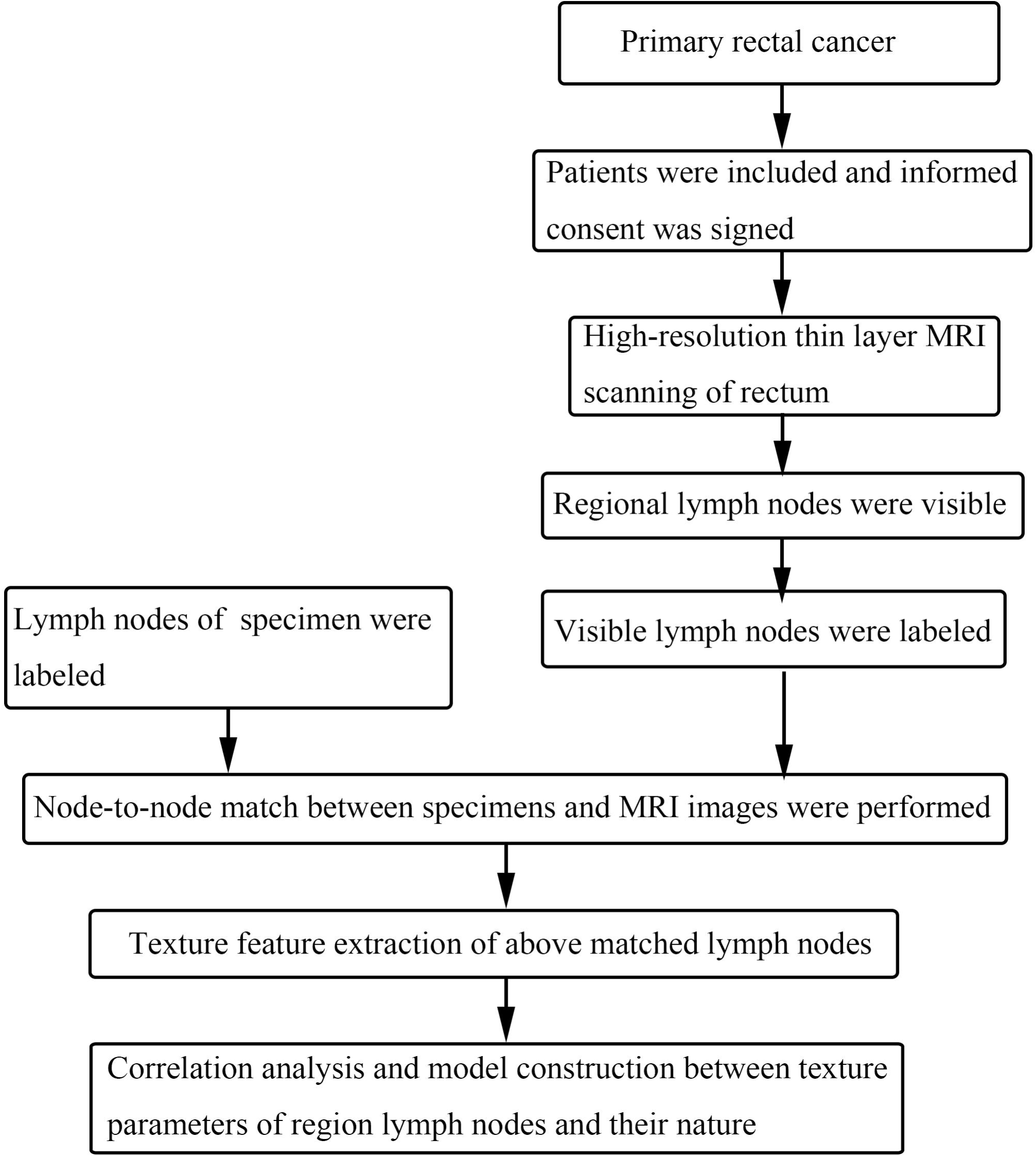
Flowchart of node-to-node match between MRI images and histopathology.

The clinicopathological data were collected for each participant, which included: age, gender, height, weight, BMI, comorbidity (hypertension, diabetes, colorectal polyps, etc.), the interval between MRI and operation (days), carcinoembryonic antigen (CEA), tumor location, tumor morphology, pathological nature of successfully matched lymph nodes, tumor differentiation, tumor, node, metastasis (TNM) staging, vessel carcinoma embolus (VCE), perineural invasion (PNI), extramural vascular invasion (EMVI), LNM in patients with rectal cancer.

### MRI Scanning Protocol

Within the 2 weeks prior to the radical operation, the patient was required to undergo a high-resolution thin-layer MRI of the rectum in our hospital. All pelvic MRI examinations were performed using a 3.0T imaging unit (Siemens Magnetom Skyro, Erlangen, Germany) with a phased-array 8-channel sensitivity encoding abdominal coil. A glycerin enema was used to clear the rectum two hours before the examination and no contrast agent was used in the rectum. The scanning sequence and related parameters included a T2 weighted imaging (T2WI) axial position (perpendicular to the diseased intestine and covering the whole pelvis), TR 4680 MS, te96 MS, matrix 384 × 281, FOV 236 mm × 260 mm, and a layer thickness of 3 mm.

### Surgery

All the eligible patients underwent radical surgery according to the principles of standard total mesorectal excision (TME) or extended TME (TME with adjacent visceral resection)(17). TME surgery is the complete removal of the rectum and the surrounding fatty envelope enclosed by the mesorectal fascia(17).

### Node-to-node Match between Specimens and MRI Images

Here, accurate one-to-one matches were ensured through the following methods: (a) TME was performed by a specialized colorectal surgeon with at least 20 years of experience in colorectal cancer surgery, which ensured a stable complete specimen collection; (b) following the operation, the chief surgeon and two radiological doctors immediately performed the node-to-node match and labeling between the specimens and MRI images; (c) the node-to-node match was based on three different sections of MRI, which included the coronal, sagittal and transverse-section; (d) the lymph nodes were eliminated following any disagreements on them(**Figure 1**).

### Region-of-Interest Segmentation and Radiomics Feature Extraction

After completing the high-resolution thin-layer MRI scan of the rectum, the MRI sequence data (stored in digital imaging and communications in medicine (DICOM)) was stored in the radiological department of the AA Hospital of AA University. The T2WI axial DICOM image was exported from the PACS system and imported into a third-party O.K (omnini dynamics, V2.08, GE) texture analysis software platform. Two radiologists, reviewed the T2WI axial image and independently marked the maximum cross-sections of rectal cancer and lymph nodes. The region of interest (ROI) was manually drawn along the lesion contour to avoid any obvious tumor necrosis, mucus area, and air. The distance between the ROI contour and the edge of the lesion was approximately 1 mm to reduce any errors caused by the partial volume effect. Overall, 56 texture parameters were extracted within each ROI, these included the first order, (12 parameters), histogram parameters (15 parameters), grey level co-occurrence matrix (GLCM) feature (7 parameters), haralick features (11 parameters), and grey level run length matrix (GLRLM) features (11 parameters) (**Table 1**). The radiomics workflow is shown in **Figure 2**.

**FIGURE 2.**
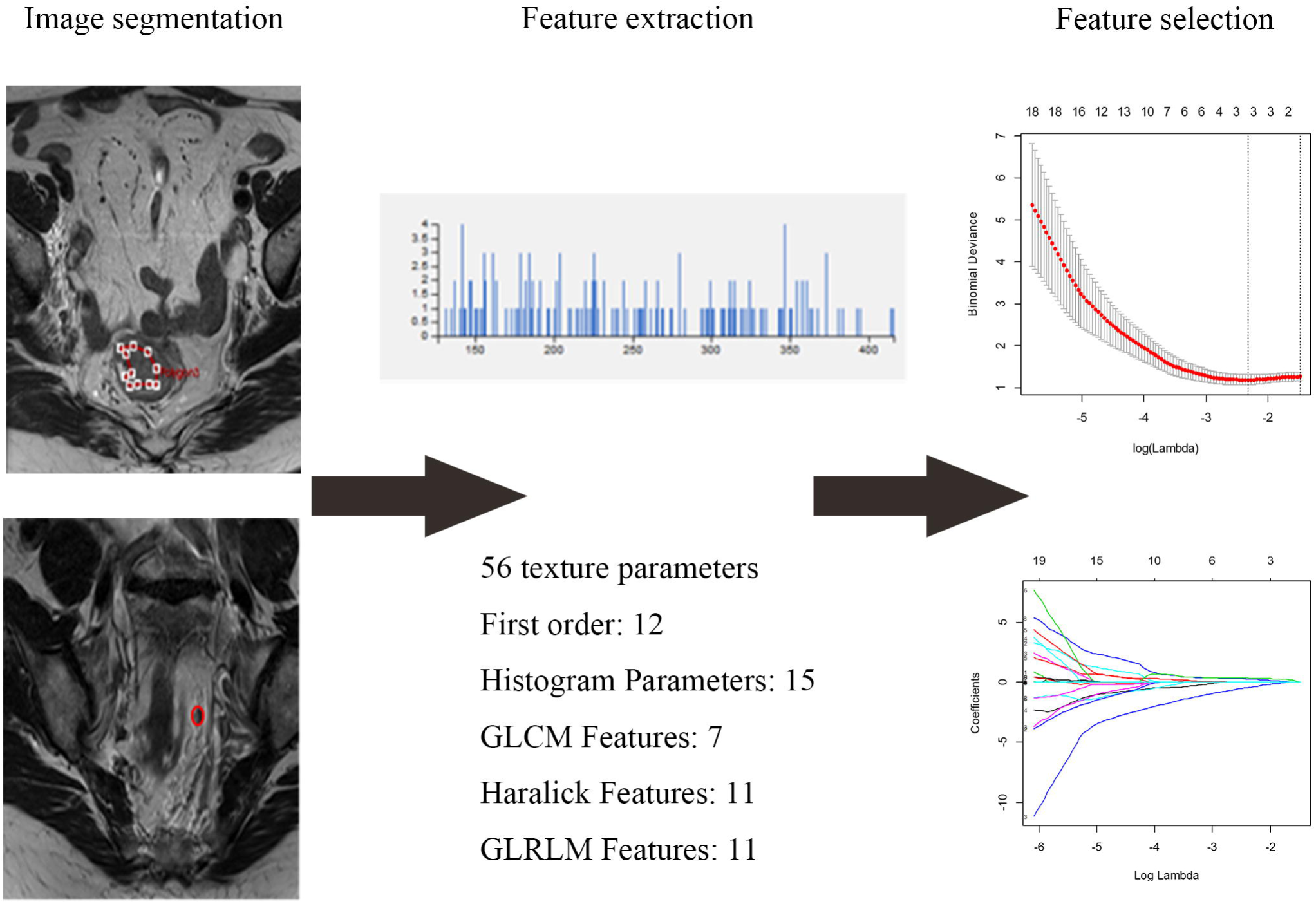
Radiomics workflow.

### Feature Selection and Radiomics Signature Development

The LASSO LR algorithm was applied to provide the further feature selection and construction of the MRI-based radiomics signature for the pretreatment prediction of LNM in rectal cancers(18). A LASSO regression model was constructed using the R language glmnet package. A 5-fold cross-validation was performed in the primary cohort to optimize the parameters of the radiomics signature. The selected features weighted by their LASSO coefficients were combined by linear combination to calculate the Rad-score of each patient, which were assessed using the AUC.

### Statistical Analysis

All statistical analyses were performed with SPSS (version 26, IBM, Armonk, NY, USA) and R software (version 4.1.2, http://www.R-project.org). The data normality was assessed by a Shapiro–Wilk test. The t-test was used for normal distribution parameters, and the Mann–Whitney U-test was used for non-normal distribution parameters. The chi-squared (χ2) or Fisher’s exact tests were used to compare the categorical variables. To obtain the independent predictors and construct the model, any items exhibiting a *P*-value < 0.1 in the univariate analysis were also included in the multivariate logistic regression analysis. A two-tailed *P*-value < 0.05 was considered statistically significant.

## RESULTS

### General Clinical Data

Here, 487 lymph nodes from 37 patients T2WI image were collected and analyzed, which included 39 metastatic lymph nodes and 11 tumor deposits. Due to no detection of relatively small lymph nodes in MRI images, only 69 lymph nodes (19 metastatic lymph nodes and 6 tumor deposits) were performed node-to-node comparisons between the radiomics and surgical lymph node histopathology. The basic clinical and pathological characteristics were shown in **Table 2**.

### The Texture Characteristics of Primary Tumors were closely related to the Tumor Differentiation

The correlation between the 56 texture features found in primary tumors with pathological characteristics was analyzed. Of the 37 patients included in this study, 20 did not possess LNM (N0) but the remaining 17 had LNM (N1-2). Likewise, 18 of the 37 patients were without PNI, while 23 patients did not possess a VCE. Finally, 8 cases were the bulge type, and 29 cases were the ulcerative type. Univariate analysis showed that patients with a VCE exhibited a lower uniformity (*P* = 0.014) and a steeper kurtosis (*P* = 0.042) compared with patients without VCE. Moreover, ulcerative rectal cancer demonstrated a lower uniformity and higher GLCM-cluster shade than the bulge type. The multivariate logistic regression analysis provided no significant difference between the various texture features with all the highlighted pathological characteristics.

Due to a lack of postoperative pathological data for a case, only data from 36 patients were included in the analysis of the correlation between primary tumor texture and tumor differentiation. This included data from 25 moderately differentiated patients (G2), 2 poorly differentiated patients (G3), and 9 moderately-poorly differentiated patients (G2-3), which were subsequently divided into two groups (a G2 group and a G2-G3 group). The univariate analysis illustrated a statistically significant difference between groups, meaning that numerous texture parameters are related to tumor differentiation. LASSO LR was used to reduce the texture features dimensions by removing the features with little correlation and redundancy. However, the three optimal features, namely the histogram - quantile5, haralick feature - difference entropy, and GLRLM-max size were obtained. The LASSO LR model was established according to these three characteristics and their weights, alongside integration of the radiology score (Rad-score), which was calculated using: Rad-score = −2.5951763036 + 0.0023059408 * quantile5 + 0.1105665700 * difference entropy + 0.0008162351 * max size. The clinical data from the two groups are shown in **Table 3**.

There was a significant difference in the Rad-score between the two groups (*P* = 0.02). The ROC curve was implemented to test the classification efficiency of the model. The results provided an AUC of 0.798, indicating that the degree of tumor differentiation in patients with rectal cancer preoperatively can be predicted from the MRI primary tumor texture parameters (**Figure 3**).

**FIGURE 3.**
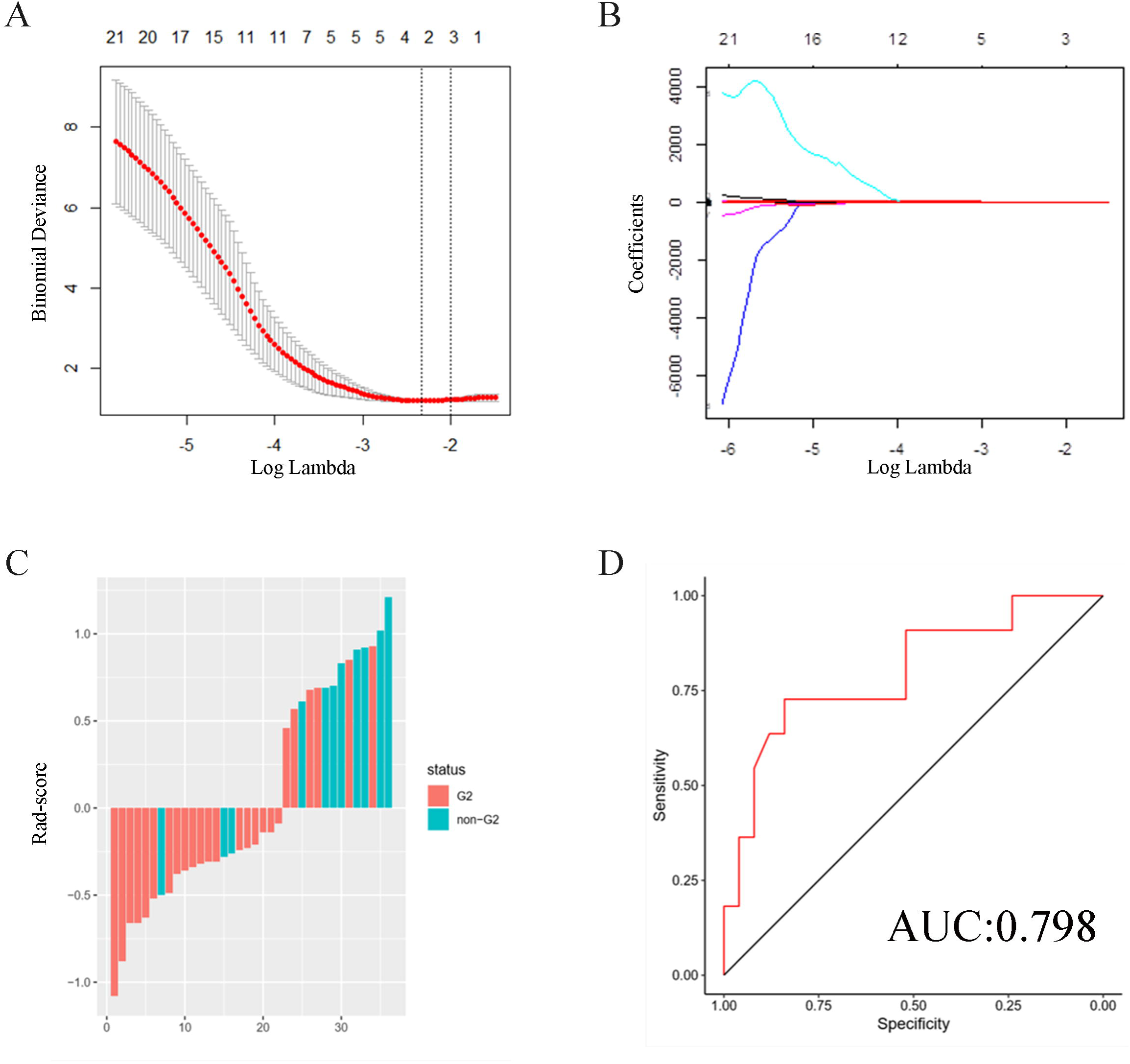
Correlation analysis between the texture characteristics of primary tumors and tumor differentiation. (A)The 5-fold cross-validation of the LASSO analysis was applied to acquire the most valuable features in predicting tumor differentiation. (B)The regression coefficients of LASSO. (C) the distribution of Rad-score in 36 patients with different degrees of differentiation. Red represents the Rad-score of moderately differentiated (G2) patients, and Blue represents the Rad-score of poorly differentiated and moderately-poorly differentiated (non-G2) patients. (D) ROC curve and AUC of lasso logistic model.

### The Texture Characteristics of Regional Lymph Node were closely related to their Nature after Node-to-node Matching

Among the 487 lymph nodes obtained in this study, 69 lymph specimens were node-to-node matched: 25 metastasis lymph specimens, including 6 tumor deposits and 19 lymph nodes. The remaining 44 nodules were nonmetastatic lymph nodes.

After taking the maximum cross-section of lymph nodes in the T2WI axial region as the ROI, a total of 56 texture features were obtained. Sixty-nine lymph nodes were randomly divided into either a training (n = 39) or verification set (n = 30). Thereafter, the 56 texture parameters are reduced in feature dimensions through lasso regression, to obtain three optimal features, which are first order-voxel value sum, haralick features-difference entropy, and GLRLM features-run-length nonuniformity (**Figure 4**). Subsequently, a lasso regression model was established:

**FIGURE 4.**
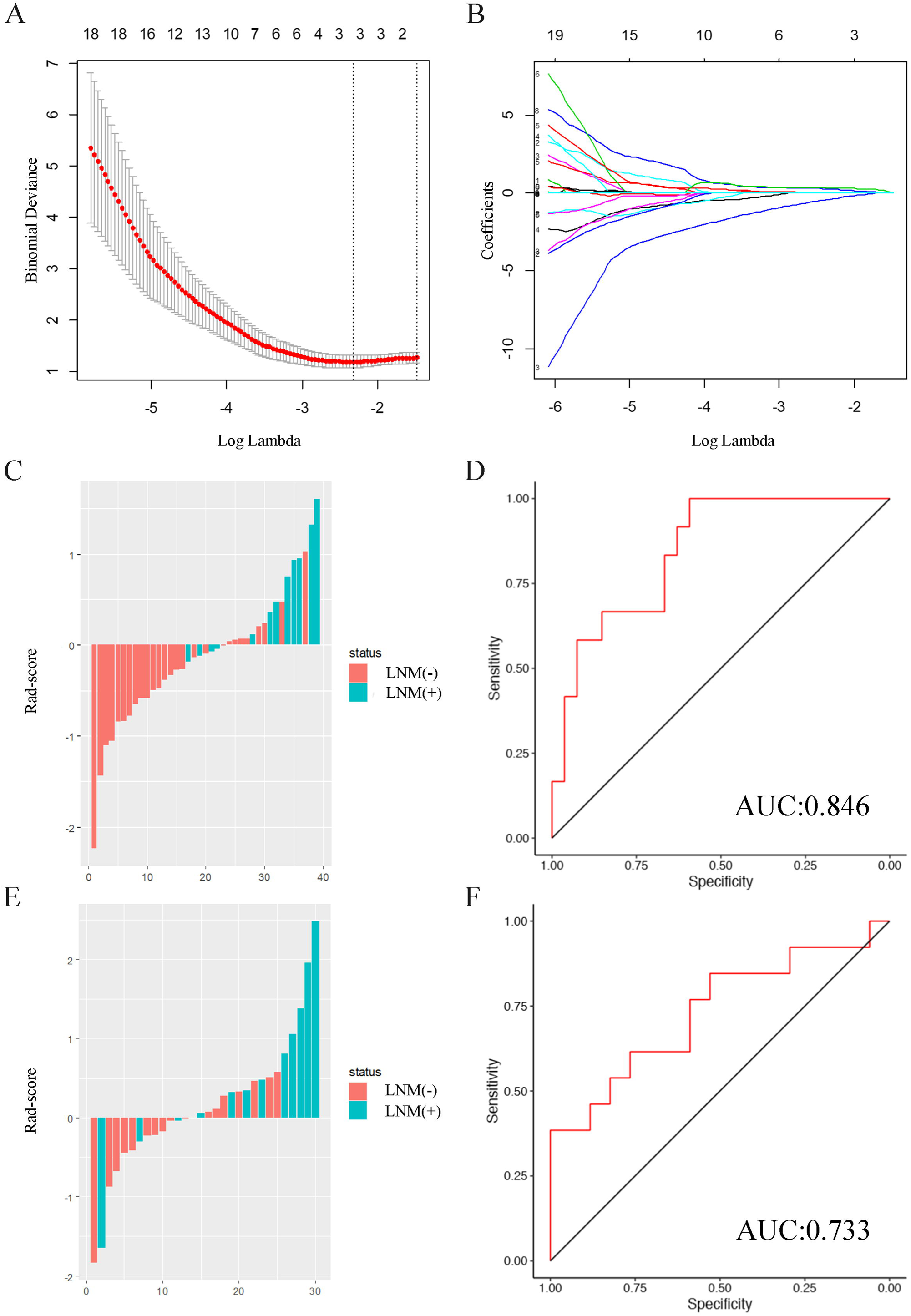
Correlation analysis between the texture characteristics of regional lymph node and their nature. (A)The 5-fold cross-validation of the LASSO analysis was applied to acquire the most valuable features in predicting LNM. (B)The regression coefficients of LASSO. (C, E) The distribution of Rad-score of 39 lymph nodes in the training and validation set. Red represents Rad-score of non-metastatic lymph nodes and blue represents Rad-score of metastatic lymph nodes. (D, F) ROC curve and AUC of lasso regression model in the training and validation set.

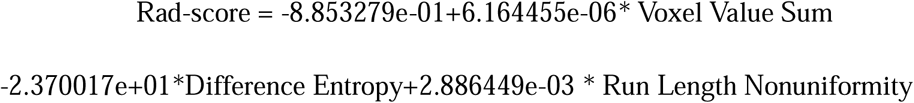

The Rad-score illustrated a significant difference between the negative lymph node and positive lymph node groups in the training set and verification sets (*P* < 0.001 and *P* = 0.032, respectively). Additionally, the ROC curve was used to test the classification efficiency of the model (**Table 4**). The results show that the AUC was equal to 0.846 in the training set and 0.733 in the verification set, indicating that the prediction model has a good classification efficiency. Furthermore, the MRI-T2WI based lymph node texture parameters can predict the nature of the regional lymph nodes of rectal cancer preoperatively.

In addition, we also found that tumor deposits had a significantly higher Rad-score than the metastatic lymph nodes (*P* = 0.042). Further, texture parameters could distinguish metastatic lymph nodes and tumor deposits (**Table 5**). The node-to-node matched lymph nodes are shown in **Figure 5-7**.

**FIGURE 5.**
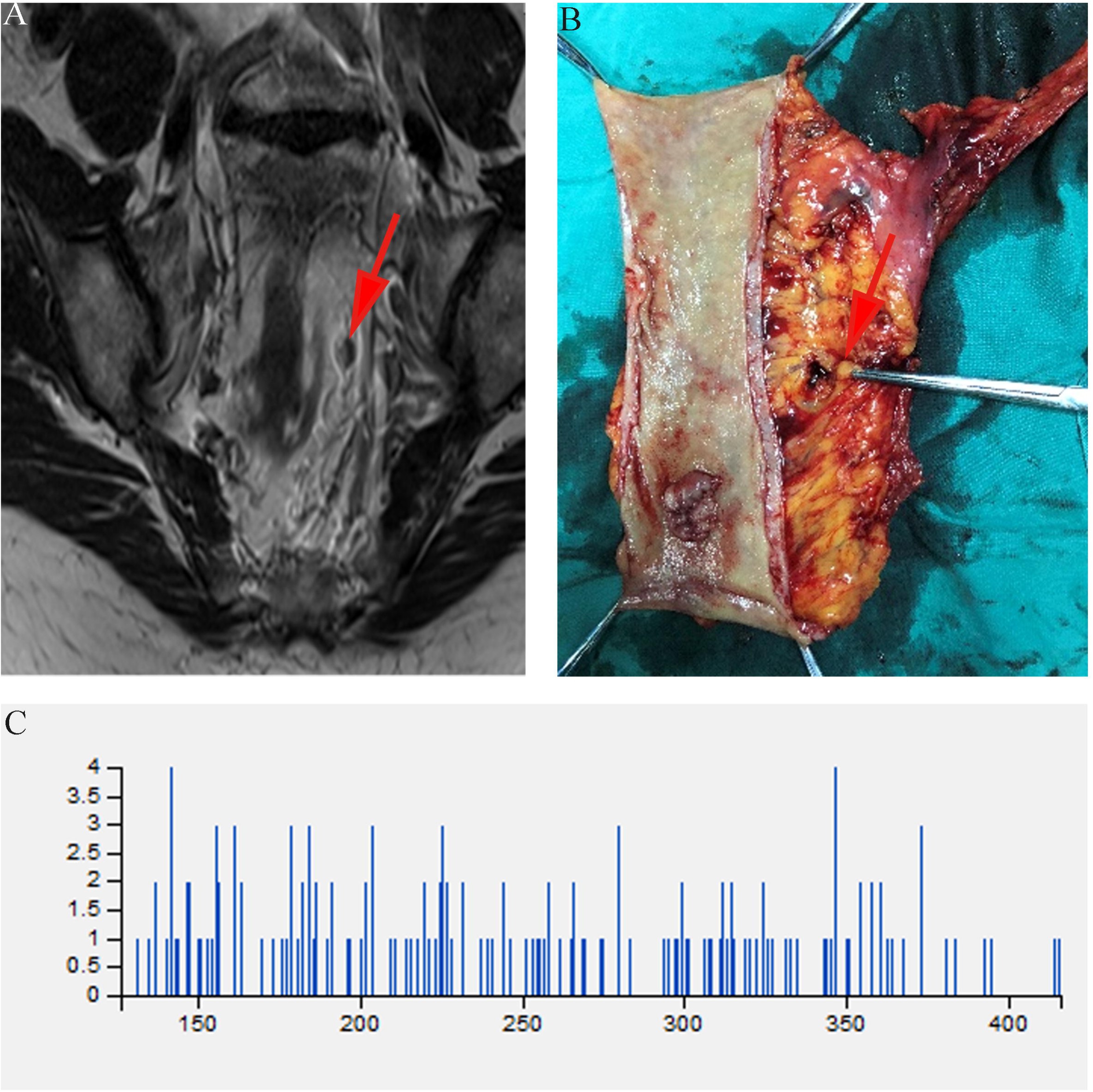
A female rectal cancer patient (age range: 60-65) was assessed to have no LNM by node-to-node method. TNM staging: pT2N0M0. Tumor morphology: bulge type. Tumor differentiation: G2(moderate differentiation). (A)Isolated lymph nodes in the mesorectum region in T2WI coronal section (red arrow); (B) The one-to-one matched lymph node detected in removed specimens (red arrow); (C) The histogram obtained from the maximum cross section of T2WI axial position of the lymph node above.

**FIGURE 6.**
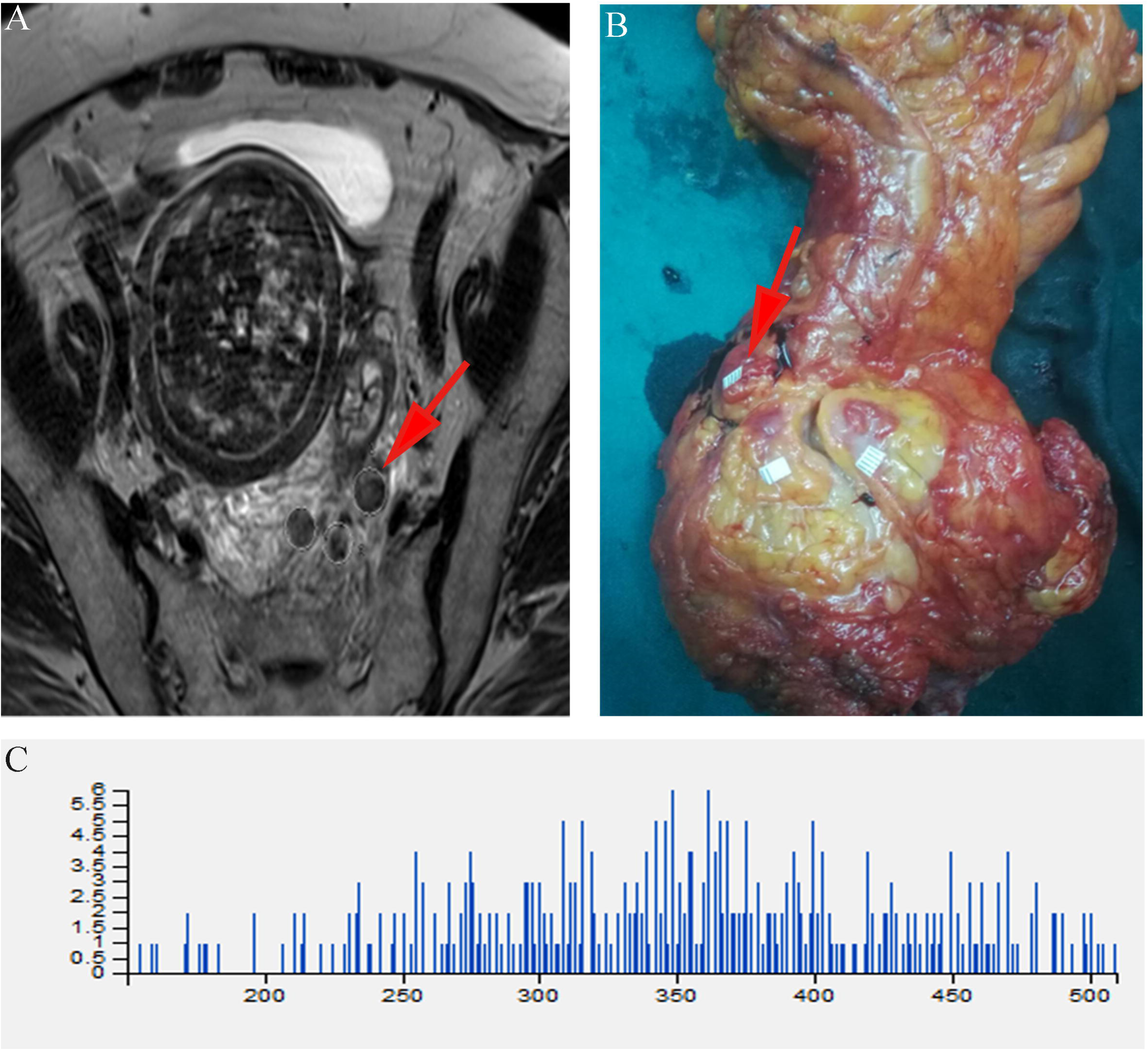
A female rectal cancer patient (age range: 66-70) was assessed to have LNM by node-to-node method. TNM staging: pT3N1bM0. Tumor morphology: ulcerative type. Tumor differentiation: G2(moderate differentiation). (A)Isolated lymph nodes in the mesorectum region in T2WI transverse section (red arrow); (B) The one-to-one matched lymph node detected in removed specimens (red arrow); (C) The histogram obtained from the maximum cross section of T2WI axial position of lymph node above.

**FIGURE 7.**
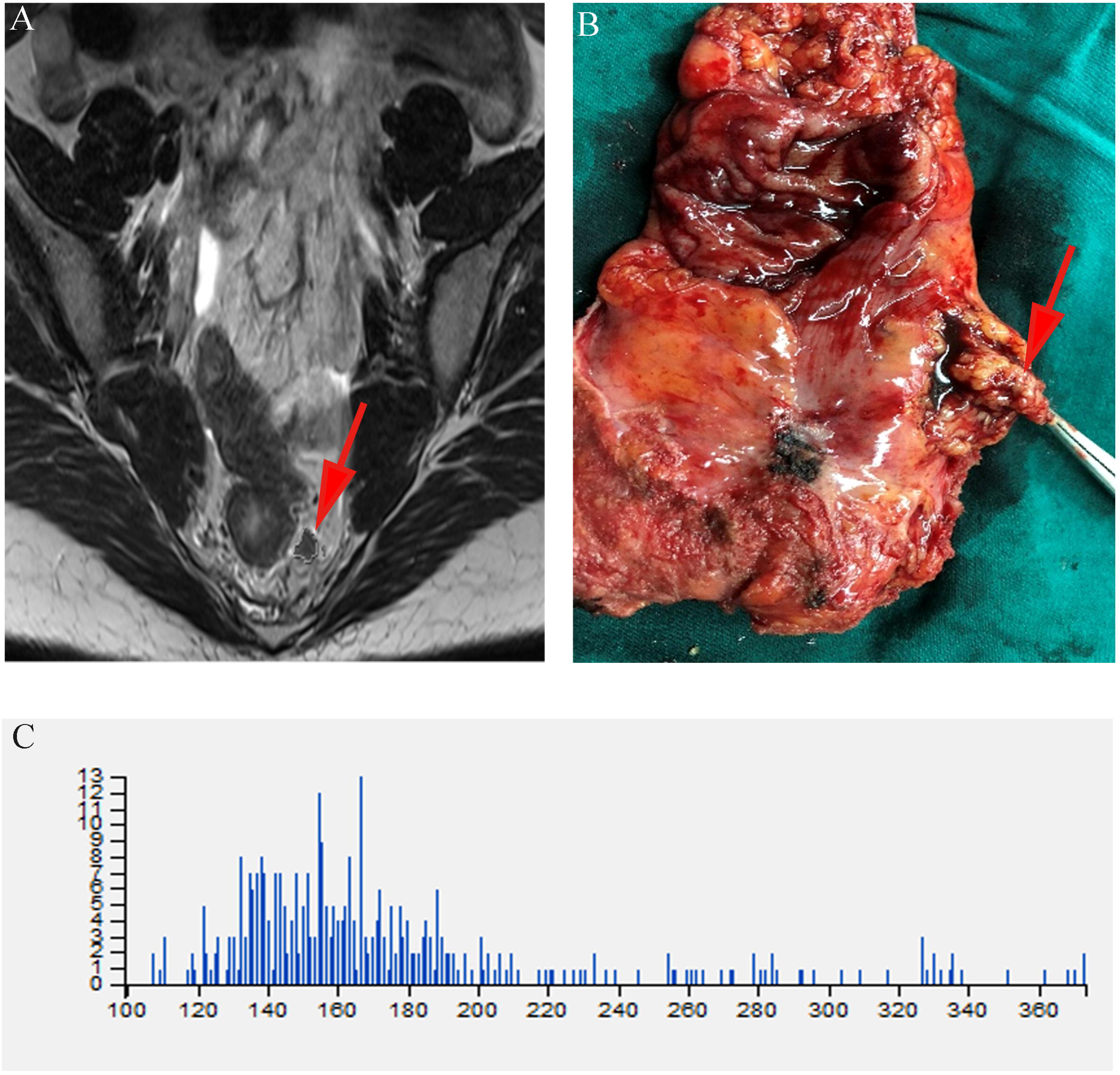
A female rectal cancer patient (age range: 36-40) was assessed to have tumor deposit by node-to-node method. TNM staging: pT2N1aM0. Tumor morphology: ulcerative type. Tumor differentiation: G2(moderate differentiation). (A)Isolated tumor deposit in the mesorectum region in T2WI transverse section (red arrow); (B) The one-to-one matched tumor deposit detected in removed specimens (red arrow); (C) The histogram obtained from the maximum cross section of T2WI axial position of tumor deposit above.

## DISCUSSION

The absence of a quantitative method able to identify LNM greatly confines the process of producing a treatment protocol for rectal cancer. In the present study, we prospectively obtained 487 lymph nodes including 39 metastatic lymph nodes and 11 tumor deposits from 37 patients with rectal cancer. Here, the relations between the primary tumors’ texture features and the subsequent pathological characteristics were analyzed. Moreover, node-to-node matching was implemented to further analyze the texture characteristics of the regional lymph nodes and their subsequent nature. We found that, the MRI textures of the primary tumors were closely related to the degree of tumor differentiation, while a validated classification model was also successfully established. After node-to-node matching between imaging and pathology, 3 texture parameters were obtained from 56 texture parameters by lasso regression, before developing a validated prediction model. The model AUC values were 0.846 in the training set and 0.733 in the validation set, demonstrating that the model can accurately identify LNM. Additionally, this study showed that the model can further distinguish metastatic lymph nodes and tumor deposits. Therefore, this model can provide reliable evidence for the early identification of regional lymph node metastasis in rectal cancer.

Texture analysis refers to a method that quantifies the pixel intensity variations by calculating the grey changes of pixels in the image, to provide the tumor heterogeneity(19). Tumors with different pathological characteristics present distinctive histological structures, and thus may also have varied texture features(20). For example, the CT texture analysis could predict tumor hypoxia and angiogenesis(19). A recent study highlighted the ability of texture analysis to visually quantify the appreciable heterogeneity by MRI. Hence, distinguishing a high-grade glioma (more heterogeneous) from a low-grade (less heterogeneous)(21). Using the established classification model in this study, the texture features of the primary tumor were demonstrated to accurately predict tumor differentiation. The accurate preoperative prediction of the tumor histological differentiation is conducive to the prediction of postoperative prognosis, alongside the formulation of a comprehensive treatment protocol. Indeed, patients presenting poorly differentiated tumors are more inclined to receive neoadjuvant treatment first. However, the sample size used for our study is small, meaning that the prediction model should be verified in future large-scale clinical studies.

Regarding the imaging diagnosis of LNM, radiomic prediction models based on the correlation analysis studies have been utilized by previous studies. Ultimately, this highlights the potential value of radiomics in this field. However, the process of retrospective designing alongside an absence of one-to-one node comparison with pathological results means their use is limited. Therefore, there is an urgent need to perform a prospective one-to-one comparison between the pathological characteristics of lymph nodes and MRI images. The difficulty in accurately identifying the suspicious lymph nodes on MRI images preoperatively, during the surgery, and on the specimens postoperatively, provides the greatest challenge for the implementation of one-to-one comparisons. Here, accurate one-to-one matches were ensured according to the methods described in the methods section. Alongside a good classification efficiency, the use of MRI—T2WI based lymph node texture parameters were found to predict the nature of regional metastatic lymph nodes in rectal cancer. The proposed quantitative model for integrated multiparametric MRI texture features could effectively help the LNM prediction and could therefore provide a novel tool to assist in the individualized management of rectal cancer. Consistently, Liu et al. conducted texture analyses of CRC primary tumor based on MRI apparent diffusion coefficient to evaluate the lymph node metastasis. However, those analyses used retrospective data, without a one-to-one comparison analysis(22). Since entropy has previously been used as an independent predictor of lymph node metastasis, our data showed that there was a significant difference in entropy between the pN0 group and the pN1-2 group(22).

Tumor deposits are described as separate tumor nodules and are usually found in the absence of lymph nodes in the pericolic or perirectal adipose tissue(23). In contrast to LNMs, tumor deposits contain unique biological features, while patients possess a poorer prognosis with them than those with a perirectal LNM(23, 24). Indeed, numerous investigations have shown that tumor deposits are linked to advanced tumor growth and higher mortality(24-29). Due to the limited evidence that lymph nodes are associated with tumor deposits, tumor deposits are predominantly distinguished from LNMs by their pathology(26). Since tumor deposits are usually discontinuous extramural extensions or focal aggregates of adenocarcinoma located in the pericolic or perirectal fat, it was unsurprising that in our study the MRI texture parameters of tumor deposits were very similar to those of rectal cancer(30). Therefore, the incorporation of texture analysis parameters to morphological findings might enhance the identification of these lesions, which could aid prognostic stratification. However, these results require further validation with a larger population in a multicenter randomized controlled trial.

Our work has two limitations. Firstly, this study is a single-center research result produced from a small sample size. Therefore, future studies should consider expanding the sample size and increasing the external verifications. Secondly, the maximum cross-sectional information of T2WI axial lymph nodes in the morphological sequence analyzed in this study did not obtain the information of the whole lymph nodes and functional MRI, such as the diffusion weighted imaging sequence. However, this study provides new research ideas and methods for use in further prospective research studies.

In conclusion, this study successfully established a classification model, which was able to predict tumor differentiation based on the primary tumors’ MRI texture. Furthermore, it successfully established an efficient MRI radiomic model, which preoperatively predicted the nature of regional lymph nodes in rectal cancer on a node-to-node match with histopathology and further distinguished between tumor deposits and metastatic lymph nodes. These two models can aid the clinical decision-making processes regarding preoperative neoadjuvant radiotherapy, chemotherapy, and surgical resection ranges. The conclusions drawn from this study need to be verified by further prospective studies with a large population.

## Data Availability

All data produced in the present study are available upon reasonable request to the authors

https://www.example.com

## DATA AVAILABILITY STATEMENT

## ETHICS STATEMENT

## AUTHOR CONTRIBUTIONS

